# Evaluating Age-Dependent Transmission and Vaccination Policy in Singapore’s SARS-CoV-2 Epidemic: A Computational Modelling Approach

**DOI:** 10.1101/2025.06.28.25330455

**Authors:** Jingyan Huang, Zhi Ling, Mousumi Roy, Shihui Jin, Jeremy WeiQuan Chan, Kelvin Bryan Tan, Swapnil Mishra, MLGH Network

## Abstract

The SARS-CoV-2 pandemic in Singapore revealed pronounced age-specific differences in disease transmission and severity. To investigate these dynamics, we developed a semi-mechanistic Bayesian hierarchical model that jointly estimated SARS-CoV-2 transmission across seven age groups from 2021 to 2023. Our model integrates both case and hospitalization data, while accounting for vaccination rollouts and time-varying contact patterns between age groups. Using a non-parametric approach, we estimated changes in contact behaviour directly from the epidemiological data, avoiding assumptions about their functional form. Results indicate that shifts in social behaviour—particularly among younger populations—significantly influenced transmission dynamics. The effective reproduction number (*R*_*t*_) for children aged 0–14 remained consistently above one, suggesting sustained transmission in this group, while reproduction number (*R*_*t*_) values for other age groups fluctuated around one with slight upward trends. The model closely matched empirical trends, with posterior estimates and 95% credible intervals aligning well with observed case and hospitalization data across all age groups. These findings underscore the critical role of age-specific contact behaviour and highlight the importance of targeted interventions during evolving pandemic phases.

## 1. Introduction

Singapore reported its first SARS-CoV-2 case in late January 2020, which was followed by sustained outbreaks in it’s foreign worker population in March/early April in Chew et al. (2020). Through various public health measures (PHMs) and contact-tracing Singapore was able to control large outbreaks in 2020. However, due to the virus mutating and exhibiting different characteristics, Singapore also experienced community transmission among its population from 2021 to 2023. The Singapore government rolled out various PHMs along with vaccination campaigns to control transmission and alleviate pressure on the healthcare system.

Age has emerged as a critical determinant of SARS-CoV-2 transmission and severity, with studies demonstrating marked heterogeneity in infection risks and outcomes across demographic groups. For instance, Monod et al. (2021) explored age-dependent variations in SARS-CoV-2 transmission in the context of the early pandemic, emphasizing the differential effects of interventions among age groups. Other notable contributions, such as those by Prem et al. (2020), examined the role of age-specific mixing patterns and their influence on the trajectory of the epidemic. Patrick G. T. Walker et al. (2020) also provided an important framework for assessing age-specific mortality and infection death rates, helping to clarify how these factors varied by demographic group. Furthermore, O’Driscoll et al. (2021) introduced a new modelling framework that integrated age-specific mortality data into pandemic projections, improving the precision of predictions on age-related disease outcomes and the effectiveness of interventions. This framework was particularly innovative in its ability to track pandemic progression in multiple age groups simultaneously, accounting for age-dependent infection rates and the varying clinical severity of the disease. Meanwhile, Yu et al. (2020) utilized generalized additive models to analyse daily new SARS-CoV-2 cases in South Korea, accounting for variations in transmission rates across different age groups. However, existing models often neglect the compounding effects of simultaneous interventions—such as vaccination and mobility restrictions—on age-specific contact patterns. These gaps persist even within age-stratified frameworks that focus on mortality and hospitalization without integrating real-time behavioural changes by O’Driscoll et al. (2021). Monod et al. (2021) further required additional sources of individual mobility data, which are seldom publicly accessible.

This study bridges the gap in the literature by providing the first non-parametric model of age-specific contact dynamics of Singapore’s SARS-CoV-2 Epidemic. Our approach leverages granular Singaporean data on case incidence, hospitalizations, and age-stratified severity. We have reconstructed the epidemiology of SARS-CoV-2 in Singapore, by seven age-specific bands, from 7 July 2021 to 30 April 2023, a period that involves persistent community driven transmission coupled with existence of waves driven by different variants of SARS-CoV-2. We extend the Bayesian statistical framework introduced by Flaxman et al. (2020) for estimating the time-varying reproduction number (*R*_*t*_) and the true number of infections. Along the lines of Davies et al. (2020); Monod et al. (2021) and O’Driscoll et al. (2021), we model age-specific SARS-CoV-2 transmission patterns from the aggregated epidemiological data. We simultaneously capture time-varying transmission intensities, *R*_*t*_, and shifts in contact patterns — encoded via autoregressive terms (AR) that reflect shifts in mobility and social behaviour. Apart from presenting results of age-specific transmission rates and true infection burden, we also present the change in contact intensities by age, which will allow us to better understand and develop targeted PHMs. These innovations allow for simultaneous evaluation of policy effectiveness while accounting for age-specific transmission patterns skewed by vaccination campaigns and other PHMs.

## 2. Data and Methods

An age-specific SARS-CoV-2 model with transmissions and interventions is investigated in this study. This section outlines general data information, contact-and-infection model with interventions, weekly contact matrix and prior for reproduction number, and the Bayesian inference framework for parameter estimation. Figure 1 shows model developed to estimate the contribution of age groups to resurgent SARS-CoV-2 epidemics in Singapore.

**Figure 1:**
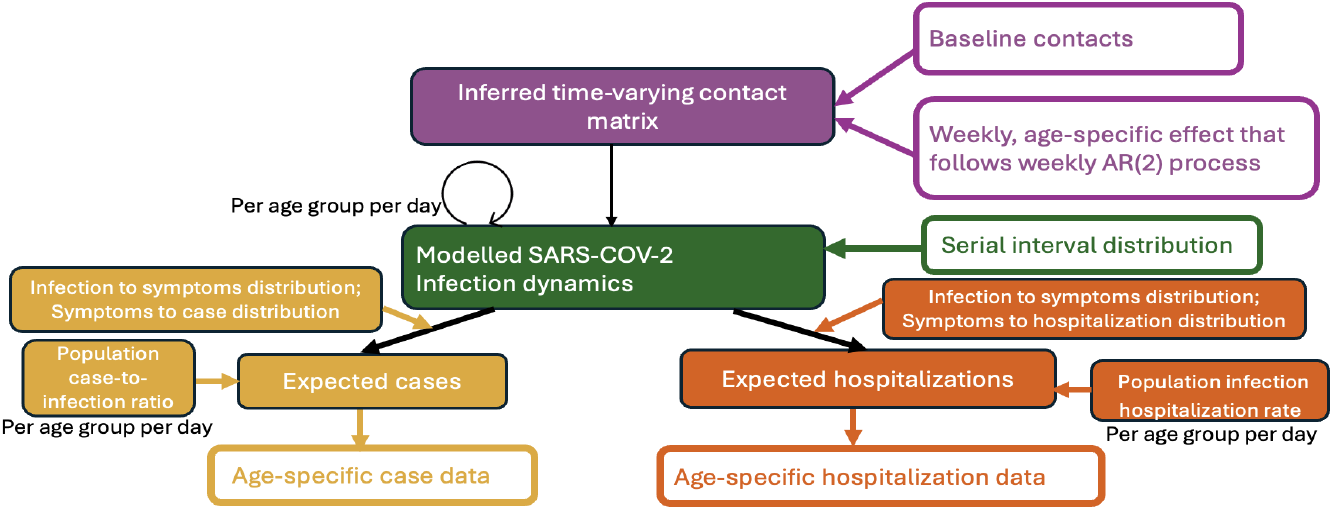
Model developed to estimate the contribution of age groups to resurgent SARS-CoV-2 epidemics in Singapore.

### 2.1. General data information

Number of new COVID-19 cases and hospitalizations reported on each day between 7 July 2021 and 30 April 2023 are gathered from Singapore’s Ministry of Health.

### 2.2. Contact-and-infection model with interventions

We denote the expected number of new infections, *i*, on day *t*, in age group *a*′ as 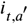. Contacts intensities are defined by the expected number of disease-relevant human contacts one person in age group *a* has with individuals in age group *a*′ in the week 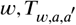, with *a, a*′ ∈ {1, 2, …, 7}. Therefore, the elements in the daily contact matrices are expressed as 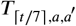. Upon contact, a proportion 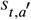 of individuals in age group on day remains susceptible to SARS-CoV-2 infection, and the susceptibility to infection with probability 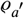. Thus, the age-specific renewal equation for infection on day *t* with weekly contact intensities is

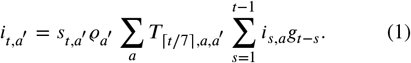

The daily generation distribution is discretized by 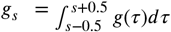 for *s* = 2, 3, … and 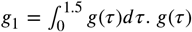 is the generation distribution which is unkn^0^own, but it can be approximated by assuming it is same as the serial interval distribution. The period we consider is from 7 July 2021 to 30 April 2023, when Delta and Omicron dominate. Based on Zeng et al. (2023), the serial interval/generation distributions of periods of Delta can be estimated to follow Gamma distribution with a mean of 4.31 days and a coefficient of variation 0.685. That is, *g*_*Delta*_ *~ Gamma*(4.31, 0.685). The serial interval of periods of Omicron is estimated to follow Gamma distributions with *g*_*Omicron*_ *~ Gamma*(2.64, 0.399).

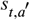 is expressed as

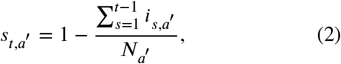

where 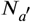 denotes the population count in age group *a*′.

The d^*a*^istribution of the time between infection and case, *π*, is approximated to be the sum of two independent random times: the incubation period (infection to onset of symptoms or infection-to-onset) and the time between onset of symptoms and recorded test (onset-to-recorded test). The infection-to-onset distribution is Gamma distributed with a mean of 5.1 days and a coefficient of variation 0.86. From Miller et al. (2022), the onset-to-recorded test distribution is Gamma distributed with a mean of 6.65 days and a coefficient of variation 0.88. The infection-to-case distribution is therefore given by:

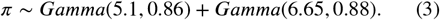

The distribution of the time between infection and hospitalization, *π*^*^, is approximated to be the sum of two independent random times: the incubation period and the time between onset of symptoms and hospitalization (onset-to-hospitalization). From Miller et al. (2022), the onset-to-hospitalization distribution is Gamma distributed with a mean of 3.88 days and a coefficient of variation 0.44. The infection-to-hospitalization distribution is therefore given by:

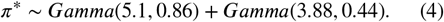

The expected number of cases 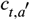, on a given day *t*, for age groups *a*′ is given by the following discrete sum:

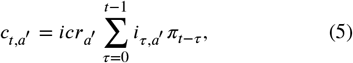

where *π* is discretized via 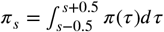 for *s* = 2, 3, … and 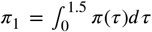 and *π*(*τ*) is the density of *π. π*_*t*−*τ*_ is the same for all age groups.

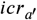 is age-specific case-to-infection ratio, which is expressed as

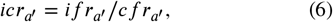

where 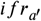 and 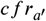 are age-specific infection fatality rate and age-specific case fatality rate respectively. *if* 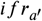 and 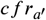 are estimated from Levin et al. (2020); Verity et al. (2020).

The daily observed cases 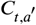 are assumed to follow a negative binomial distribution with mean 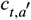 and variance 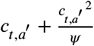, where *ψ* follows a half normal distribution, i.e.

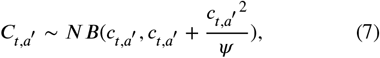

with *ψ ~ N*^+^(0, 5).

The expected number of hospitalization data 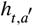, on a given day *t*, for age groups *a*′ is given by the following discrete sum:

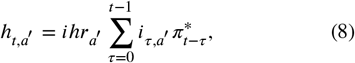

where 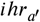 is age-specific infection hospitalization rate. 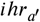 is estimated from Hozé et al. (2021).

The daily observed hospitalization counts 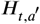 are assumed to follow a negative binomial distribution with mean 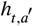 and variance 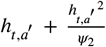, where *ψ*_2_ follows a half normal distribution, i.e.

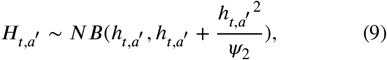

with *ψ*_2_ *~ N*^+^(0, 1). From equation (1), the time-varying reproduction number on day *t* from one infectious person in age band *a*′ is expressed as

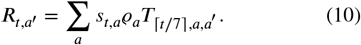

Next, we consider how the age-specific SARS-CoV-2 model incorporates the intervention of vaccination regimen. Singapore government mandated that booster doses be administered within six months after the second dose, leading to a high booster uptake among the population. Accordingly, in our analysis, we assumed no vaccine protection before July 2022 and vaccine effectiveness stayed the same throughout the ten months between July 2022 and April 2023. 30 June 2022 was chosen since 96% of the eligible populace had completed their vaccination regimen as of June 2022. We set *T*_*vaccine*_ = 303, which refers to total days throughout the ten months. After *T*_*vaccine*_, 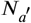 is reduced so that susceptible size of people, 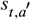, in (2) is reduced. 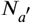 is reduced according to the follows,

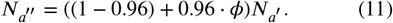

The variable *ϕ* is modelled as a normal distribution, *ϕ ~ N*(0.8, 0.05). This is based on the finding that 96% of the eligible population had completed the vaccination regimen and were perceived to have an 80.3% susceptibility to SARS-CoV-2, according to the parsimonious classifier described in Raza et al. (2022). Consequently, the mean of *ϕ* is approximated as 0.8.

### 2.3. Weekly contact matrix and prior for reproduction number

Seven age bands include [0 − 14], [15 − 29], [30 − 39], [40 − 49], [50 − 59], [60 − 69], [70+] and the length of time is 602 for period between 1 September 2021 and 30 April 2023. The population counts *N*_*a*_ in each age group are expressed as *N*_1_ = 581.7, *N*_2_ = 716.54, *N*_3_ = 623.67, *N*_4_ = 612.69, *N*_5_ = 603.04, *N*_6_ = 552.83, *N*_7_ = 458.81 in thousands. *N* is the summation of *N*_*a*_ across all age groups.

The prior distribution for initial value of reproduction number, *R*_0_, was chosen to be

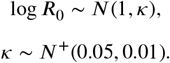

Let *AvgCntct* denote average contact across all age groups. *AvgCntct* is calculated by contact matrix 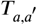 and population by age.

The weekly contact matrix is derived from Monod et al. (2021), which is shown as

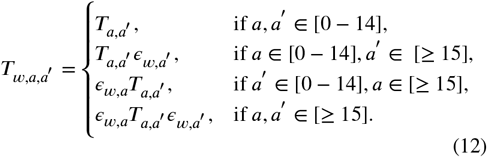

where the contact matrix 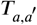 is from R package “squire” by fixing “country code = Singapore” in Watson et al. (2021); Patrick G. T. Walker et al. (2020). We model *ϵ*_*w,a*_ as weekly, age-specific effect and follow the weekly AR(2) process, as shown in Unwin et al. (2020). Assume that

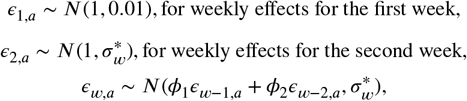

for *w* = 3, 4, 5, …. The prior for *ϕ*_1_ is a *N*(0.8, 0.05) distribution and the prior for *ϕ*_2_ is a *N*(0.1, 0.05) distribution. The prior for *σ*_*w*_, the standard deviation of the stationary distribution of *ϵ*_*w*_ is chosen as

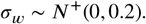

The standard deviation of the weekly updates to achieve this standard deviation of the stationary distribution is

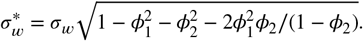

We consider the initial values of number of cases and from equation (6) priors for parameters of initial values of number of infections *τ* are assumed that

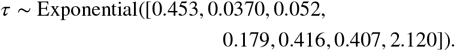

We have

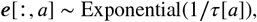

and

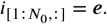

Consider periods of initial values of infections *N*_0_ = 14 and seeded period of 112 days since longer period of initial values of infections and seeded period make *R*_*t,a*_ more stablized when starting to estimate 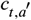 and 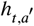.

### 2.4. Bayesian inference

Contact-and-infection model was fitted via probabilistic programming language Stan (version 2.33.1) by Carpenter et al. (2017), using the No-U-Turn Sampler (NUTS). CmdStan programs were invoked using CmdStanR Interface (version 0.6.1) by Axen et al. (2023). 4 chains were run in parallel for 1,500 iterations, of which the first 500 iterations were specified as warm-up. There were no divergent transitions after warmup. R and Stan codes are available at https://github.com/mlgh-sg/Age-Specific-SARS-CoV-2/tree/main/R_Stan_Code.

## 3. Results

We fit the age-specific SARS-CoV-2 model with transmissions and interventions in Section 2 to Singapore agespecific SARS-CoV-2 case and hospitalization data from 7 July 2021 until 30 April 2023. The results of model fits and key generated quantities for age group [0-14], [15-29], [30-39], [40-49], [50-59] [60-69], [70+], are shown in this Section. The left columns in Figure 2 and 3 show the estimated age-specific effective reproduction number, posterior mean estimate with 95% credible intervals. The middle column shows the estimated age-specific hospitalization data, posterior mean estimate with 95% credible intervals. The right column shows the estimated age-specific case data, posterior mean estimate with 95% credible intervals. The added intervention such as mass vaccination with transmission-blocking vaccines reduced the population’s susceptibility starting from 1 July 2022 since 96% of the eligible population had completed their vaccination regimen by June, 2022. We estimate that *R*_*t*_ exceeds one only for the youngest age group [0-14] while fluctuating around one with slight increasing trends for age group [40+]. For the [0–14] age group, *R*_*t*_ hovers around 1.3 prior to September 2022, followed by an upward trend thereafter. For age groups 40 and above, the *R*_*t*_ value ranges between 0.5 and 1.5 from 31 January 2022 to 30 April 2023. Figure 3(H) indicates *R*_*t*_ value ranges between 0.7 and 1.5 during the whole study period. The model closely matches empirical trends, with posterior estimates and 95% credible intervals aligning well with observed case and hospitalization data without age stratification.

**Figure 2:**
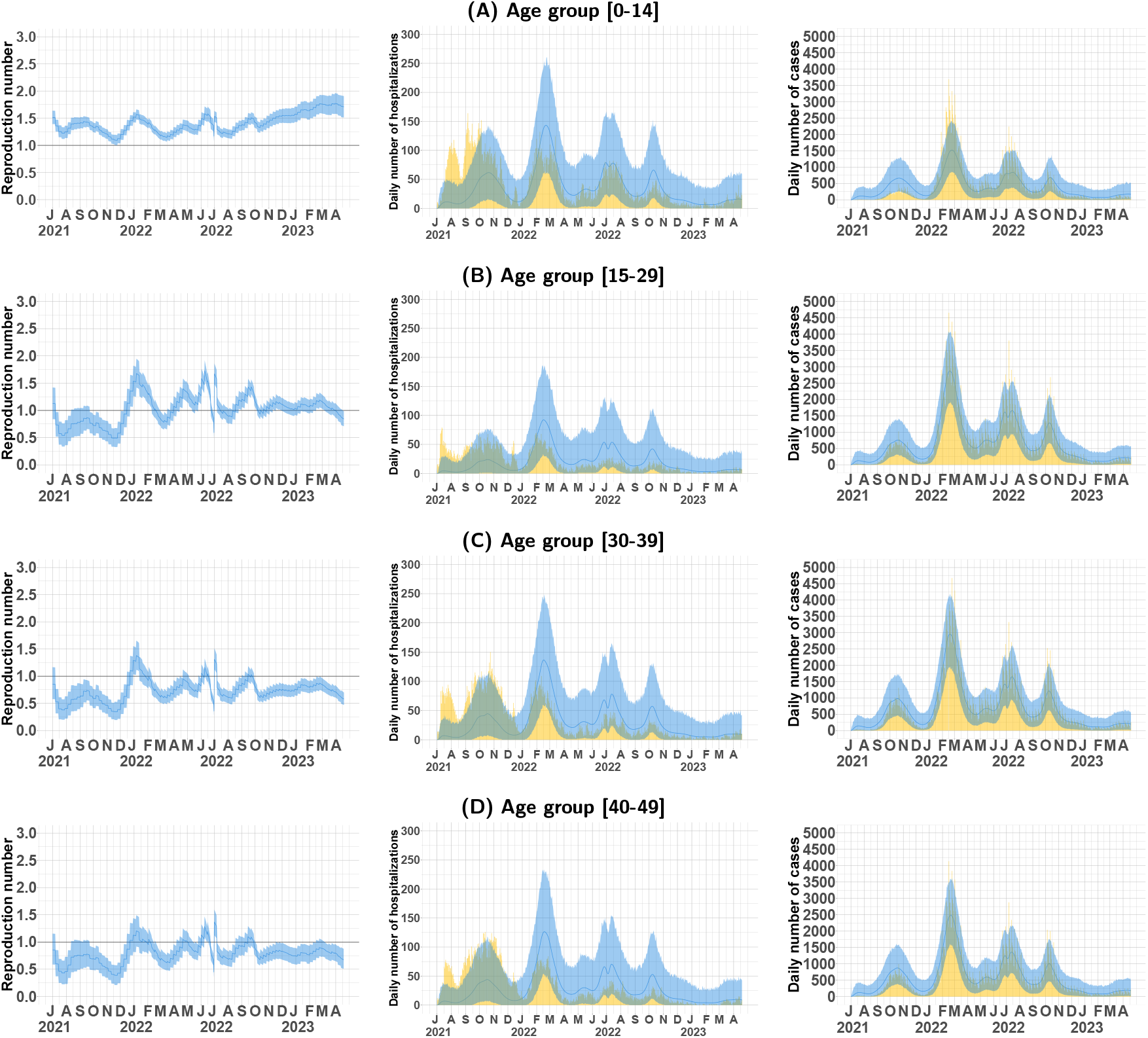
Model fits and key generated quantities for age groups [0–14], [15–29], [30–39], and [40–49]. (Left) Estimated age-specific effective reproduction number, posterior mean estimate (dark blue line), and 95% credible intervals (light blue ribbon). (Middle) Observed daily age-specific SARS-CoV-2 hospitalization data (yellow barplot) versus posterior mean estimate (dark blue line) and 95% credible intervals (light blue ribbon). (Right) Observed daily age-specific SARS-CoV-2 case data (yellow barplot) versus posterior mean estimate (dark blue line) and 95% credible intervals (light blue ribbon).

**Figure 3:**
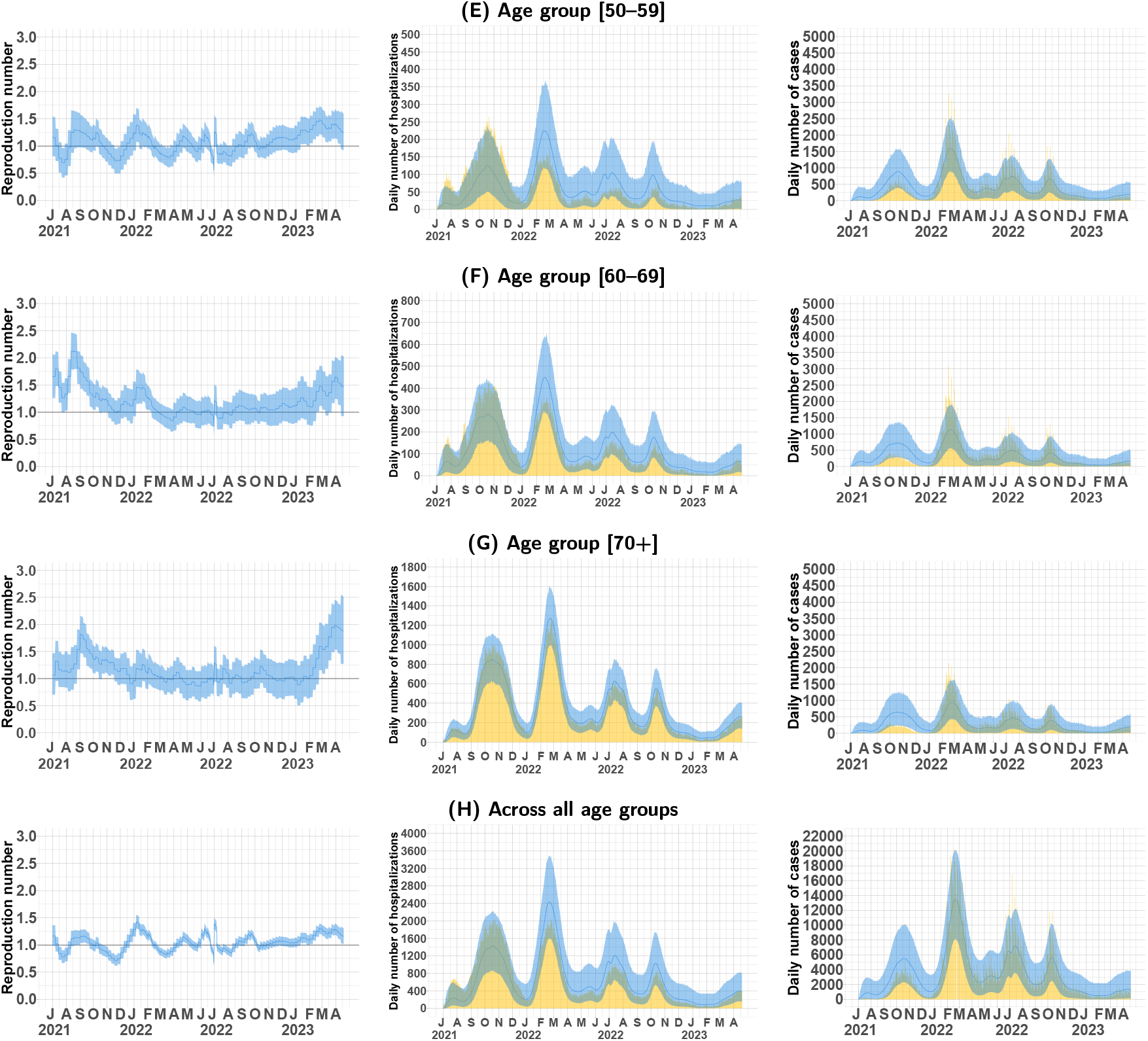
Model fits and key generated quantities for age groups [50–59], [60–69], [70+], and across all age groups. (Left) Estimated age-specific effective reproduction number, posterior mean estimate (dark blue line), and 95% credible intervals (light blue ribbon). (Middle) Observed daily age-specific SARS-CoV-2 hospitalization data (yellow barplot) versus posterior mean estimate (dark blue line) and 95% credible intervals (light blue ribbon). (Right) Observed daily age-specific SARS-CoV-2 case data (yellow barplot) versus posterior mean estimate (dark blue line) and 95% credible intervals (light blue ribbon).

Estimated SARS-CoV-2 infections (cases) before / after intervention of vaccine, Singapore average from 7 July 2021 until 30 April 2023 and shares in population for each age group are shown in Figure 4. The estimated proportions of SARS-CoV-2 infection for the age group [50+] reduce, possibly due to the significant protective effect of the vaccine; while the estimated proportions of SARS-CoV-2 infection for the age group [15-39] increase. Reported case proportions increase for age group [15-29] while decrease for age group [30-39] after intervention of vaccine. Reported case proportions for other age groups rarely changed from 7 July 2021 to 30 April 2023. The relative attack rates in age group [0-14], [15-29], and [30-39] estimated from infections decrease compared to the estimation from cases; while for the age group [50-59], [60-69] and [70+], the trend is vice versa.

**Figure 4:**
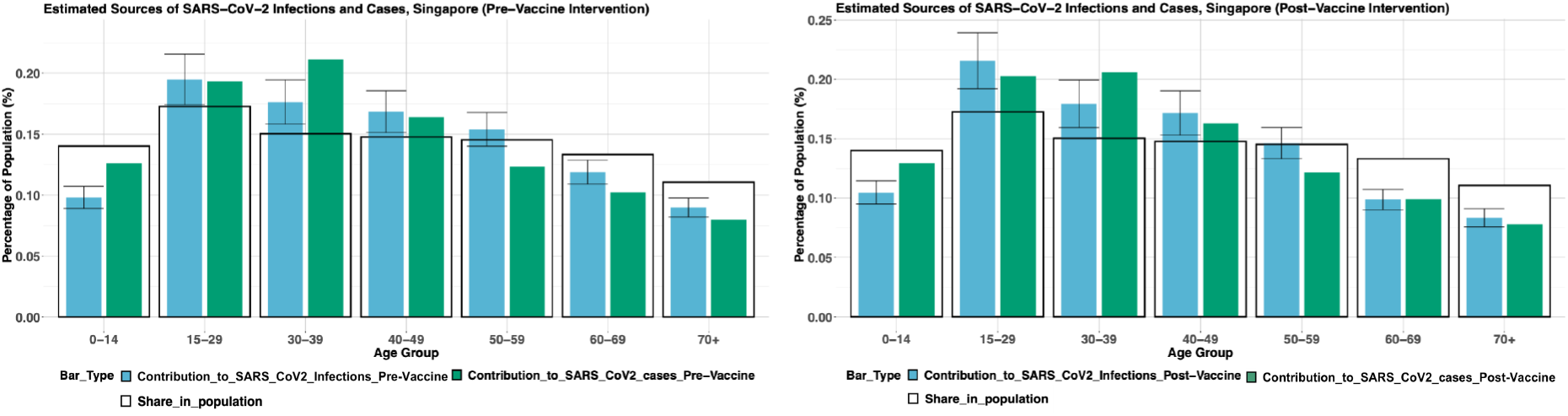
Comparison of SARS-CoV-2 estimated infections and true cases Pre-Vaccine (before intervention of vaccine) and Post-Vaccine (after intervention of vaccine) in Singapore. The proportion of SARS-CoV-2 estimated infections in each age group to all estimated infections (blue barplot) with the proportion of SARS-CoV-2 true cases in each age group to all true cases (green barplot) and the proportion of population in each age group to the total population (blank barplot). Data from 7 July 2021 to 30 April 2023.

Figures 9-15 in the supplementary note illustrate a comparison of SARS-CoV-2 infection proportions for different age groups in Singapore, both before and after the introduction of the vaccine. For age groups [0-14], [15-29], [30-39] and [40-49], proportions of infections caused by age groups [0-39] increase while infections caused by age groups [50+] decrease. For age groups [50-59], proportions of infections caused by age groups [0-39] decrease while infections caused by age groups [50+] increase. For age groups [70+], proportions of infections caused by age groups [0-49] decrease while infections caused by age groups [60+] increase. For age group [50-59] and [70+], their social activities with similar age groups especially [50+] and [60+], increase after the vaccination. Hence, proportions of infections caused by [50+] and [60+] increase.

The google mobility trends play a limited role in identifying the age-specific behavioural drivers contributing to SARS-CoV-2 transmission, as shown in Figure 5(A). The detailed, age-specific SARS-CoV-2 hospitalization data and case data are used to estimate how interventions, changes in contact intensities, age, and other factors have interacted and led to resurgent disease spread. Figure 5(B) shows the contact matrices for the time before rise, before fall, before rise, before interventions of vaccine, before rise and before rise of observed daily SARS-CoV-2 hospitalization or case data. The average contact intensities of the contact matrices in the weeks of 9 February 2022, 29 June 2022, 13 July 2022 and 5 October 2022 were greater than one, suggesting that the increasing contact intensities likely accelerated the transmission of SARS-CoV-2 and contributed to the observed patterns in SARS-CoV-2 case and hospitalization rates.

**Figure 5:**
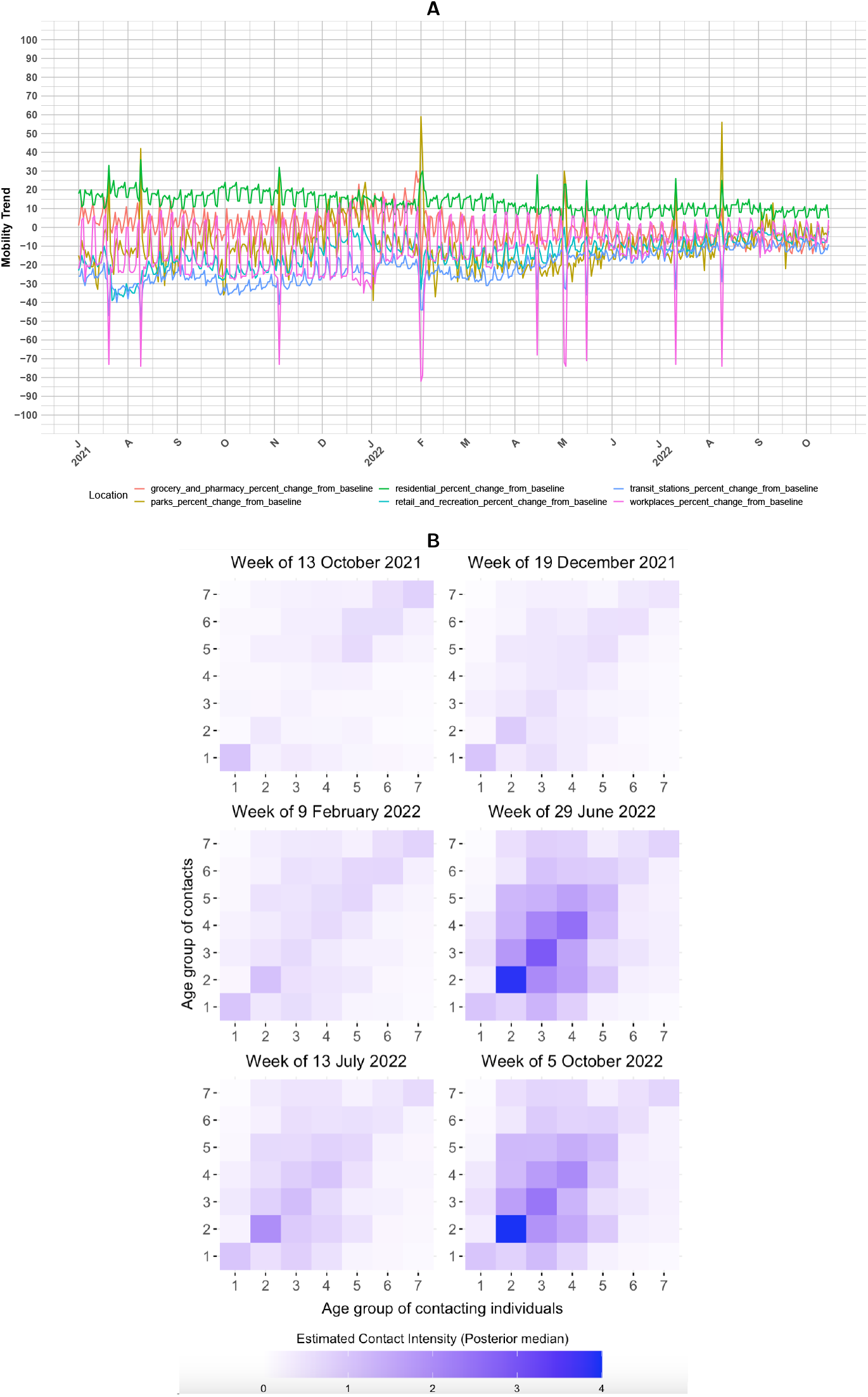
(A) Google Mobility trends across different locations (retail and recreation, grocery and pharmacy, residential, transit stations, parks, and workplaces) in Singapore. This figure illustrates how visits and length of stay at these places change relative to a baseline, where the baseline is the median value for the corresponding day of the week, for example, during the five-week period from 3 January to 6 February 2020. (B) Contact matrices for weeks of 13 October 2021, 19 December 2021, 9 February 2022, 29 June 2022, 13 July 2022, 5 October 2022 that align with time before rise, before fall, before rise, before interventions <u>of vaccine, before rise and before rise of observed daily SARS-CoV-2 hospitalization or case data.</u>

Sensitivity analyses were conducted to assess central modelling assumptions on the standard deviations in *ihr* noise, standard deviations in *ψ*_2_ (a parameter in equation (9)), and are reported in Table S1, Table S2, Figure 1-8 in the supplementary note. Our findings on the age groups that drive SARS-CoV-2 transmission were found to be robust to these assumptions.

## 4. Discussion

In this study, we jointly model the Singapore SARS-CoV-2 epidemic at the age-level, using age-specific SARS-CoV-2 case and hospitalization data within a semi-mechanistic Bayesian hierarchical framework for SARS-CoV-2 in Singapore. Our model incorporates vaccination as a key intervention and estimates the number of cases, hospitalizations, and the time-varying reproduction number (*R*_*t*_) for each age group from 7 July 2021 until 30 April 2023. One innovation of this study is the explanation of age-specific transmission dynamics using a semi-mechanistic Bayesian hierarchical framework. Another key innovation is the estimation of age-specific reproduction numbers without relying on mobility trends, which are unavailable in some Southeast Asian countries.

The posterior mean estimate with 95% credible intervals Fit the age-specific data well for both hospitalization data and case data. The added intervention such as mass vaccination with transmission-blocking vaccines reduced the population’s susceptibility starting from 1 July 2022 since 96% of the eligible population had completed their vaccination regimen by June, 2022. The age-specific reproduction numbers from July 2022 indicate that mass vaccination could help bring the reproduction number and resurgent SARS-CoV-2 epidemics under control. We estimate that *R*_*t*_ exceeds one only for the youngest age group [0-14] while fluctuating around one with slight increasing trends for age group [40+]. For the [0–14] age group, *R*_*t*_ hovers around 1.3 prior to September 2022, followed by an upward trend thereafter. For this age band, *R*_*t*_ reaches local maximum on 15 July 2022. By 15 July 2022, Singapore had eased several COVID-19 restrictions, including lifting of group size limits, reopening of schools and preschools and relaxation of mask mandates. reaches local minimum near December 2021 and 10 August 2022. In December 2021, Singapore extended its national vaccination programme to include children aged 5 to 11 years, marking a significant step in controlling COVID-19 transmission among younger populations. By September 2022, Singapore had stabilized its COVID-19 situation through continued vaccination efforts and PHMs. Ongoing vaccination efforts focus on immunizing children aged 5 to 11 and providing booster doses to eligible individuals. For age groups 40 and above, the *R*_*t*_ value ranges between 0.5 and 1.5 from 31 January 2022 to 30 April 2023, suggesting relatively stable transmission without significant acceleration or decline. Instead of mobility trend, the age-specific reproduction numbers in Singapore are estimated by weekly effects based on AR(2) process. The estimation of age-specific reproduction numbers without mobility trend in Singapore is non-trivial since there are no mobility trends in some Southeast Asian countries, like Philippines, Indonesia, Myanmar, and so on.

Right columns in Figure 2 and Figure 4 provide evidences that SARS-CoV-2 in Singapore from 7 July 2021 until 30 April 2023 have been driven by people in age group [15-49], in particular, people aged 15 to 39, before and after intervention of vaccine. People aged 15 to 49 accounted, after intervention of vaccine in July 2022, for an estimated 57% of SARS-CoV-2 infections in Singapore, with only 10% originated from age group [0-14]. Whereas, people aged 35 to 49 accounted, after school reopening in October 2020, for an estimated 72.2% of SARS-CoV-2 infections in the US locations considered in Monod et al. (2021). Over time, the share of age groups among reported cases has been constant, suggesting that young adults are unlikely to have been contributing to ongoing transmission and primary source of SARS-CoV-2 epidemics and that, instead, changes in behaviour among the broader group aged 15 to 49 underlie resurgent SARS-CoV-2 in Singapore from July 2021 until April 2023. This study indicates that in Singapore where novel, highly transmissible SARS-CoV-2 lineages have not yet become established, additional interventions among adults aged 15 to 49, such as mass vaccination with transmission-blocking vaccines, testing or social distancing, could bring resurgent SARS-CoV-2 epidemics under control. This aligns with intervention measures among adults aged 20 to 49 shown in Monod et al. (2021).

Supplementary notes illustrate a comparison of SARS-CoV-2 infection proportions for different age groups in Singapore, both before and after the introduction of the vaccine. For age groups [0-14], [15-29], [30-39] and [40-49], proportions of infections caused by age groups [0-39] increase while infections caused by age groups [50+] decrease. The reason is that SARS-CoV-2 vaccination programme in Singapore is available for people aged 12 and above on 31 May 2021. Therefore, vaccination programme in Singapore protects people aged 12 and above, especially for middle-age and elderly people.

The google mobility trends indicate a stable visits and length of stay at these places change to a baseline for all different locations in Singapore. These google mobility trends play a small role in identifying signatures of age-specific, behavioural drivers of SARS-CoV-2 transmission as they are relatively stationary across all locations. The contact matrices for the time before rise, before fall, before rise, before interventions of vaccine, before rise and before rise of observed daily SARS-CoV-2 hospitalization or case data are shown. The average contact intensities for contact matrices for week of 9 February 2022, 29 June 2022, 13 July 2022, 5 October 2022 exceeded one, suggesting that the increasing contact intensities likely accelerated the transmission of SARS-CoV-2 and contributed to the observed patterns in SARS-CoV-2 case and hospitalization rates.

One of the limitations of this study is the restriction in analysis time frame. The age-specific SARS-CoV-2 model was calibrated using Singapore’s SARS-CoV-2 data from 7 July 2021 to 30 April 2023. Future work could expand this model to alternative time periods for a wider applicability to other countries. Another limitation is the assumption of constant case-ascertainment rate during the study period.

Further research could offer valuable information by developing SARS-CoV-2 models that account for both age-specific and country-specific effects. Furthermore, exploring models that incorporate varying intervention strategies in different countries can improve our understanding of how different policies influence the dynamics and results of SARS-CoV-2 transmission.

## Data Availability

All data produced in the present study are confidential

