## Supplementary Note for "Evaluating Age-Dependent Transmission and Vaccination Policy in Singapore’s SARS-CoV-2 Epidemic: A Computational Modelling Approach"

#### 1. Sensitivity analysis

Sensitivity analysis reveals the extent to which results depend on uncertain parameters and modelling choices, and can diagnose model misspecification. We vary some components of our model and recompute the model fits and key generated quantities (including estimated age-specific effective reproduction number, estimated age-specific case data and estimated age-specific hospitalization data) for varied age groups. Overall, we perform the sensitivity analyses for varied standard deviations in $ihr\_noise$ and $\psi_2$, which is a parameter in equation (9) in main document. Supplementary Table S1 summarizes our sensitivity analyses and their categories.

| Category Name | Analyses |
|---|---|
| Standard deviation in $ihr\_noise$ and standard deviation in $\psi_2$ | $ihr\_noise \sim N(1, 0.1); \psi_2 \sim N(0, 1)$<br>$ihr\_noise \sim N(1, 0.3); \psi_2 \sim N(0, 2)$<br>$ihr\_noise \sim N(1, 0.8); \psi_2 \sim N(0, 1)$<br>$ihr\_noise \sim N(1, 1); \psi_2 \sim N(0, 0.1)$ |

**Table S1:** Sensitivity analyses and their categories.

*Corresponding author
 (J. Huang); (S. Mishra)
https://mlgh-sg.com (S. Mishra)
ORCID(s): 0000-0002-8759-5902 (S. Mishra)

##### 1.1 $ihr\_noise \sim N(1, 0.1); \psi_2 \sim N(0, 1)$

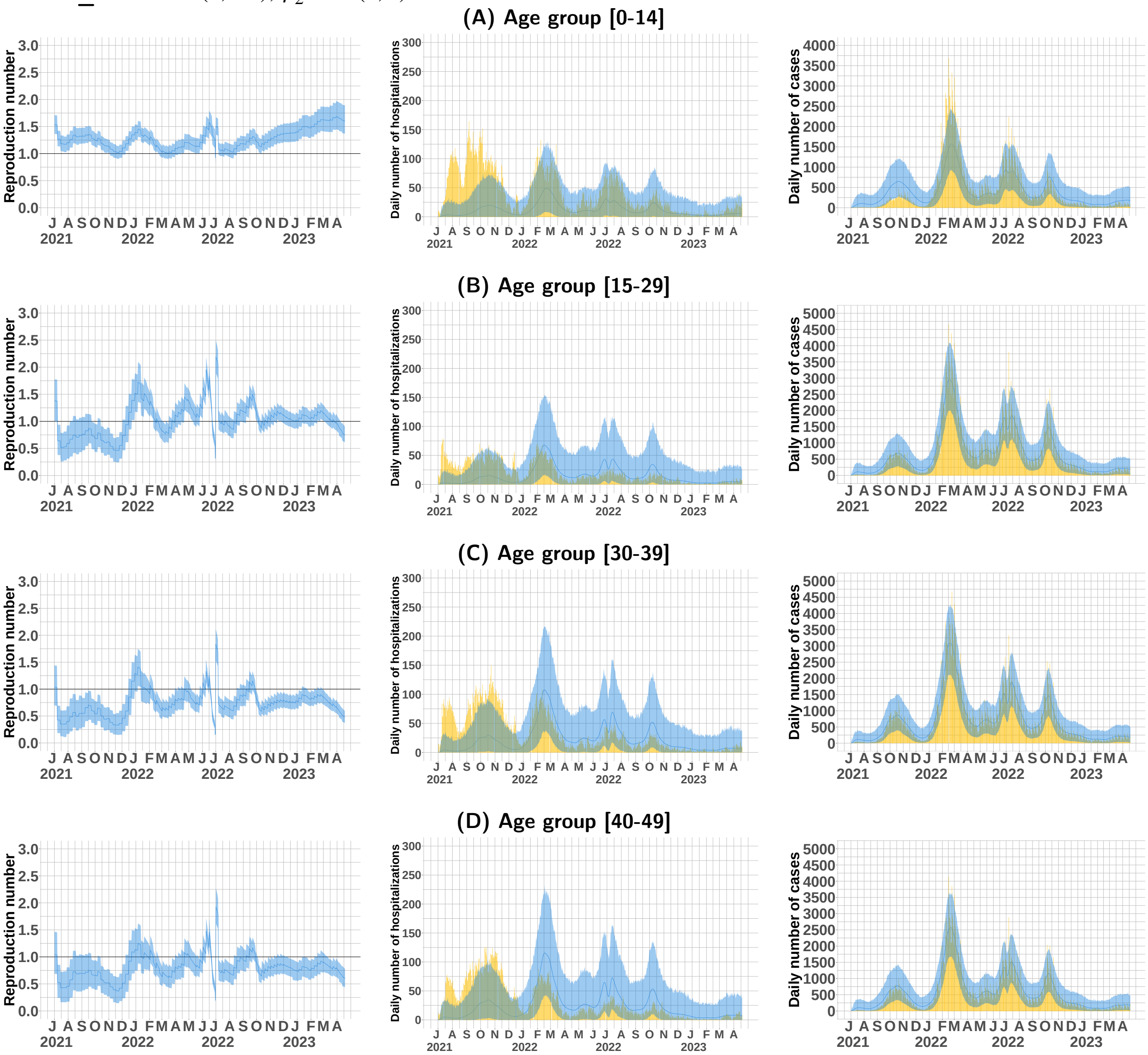


**Figure 1:** Model fits and key generated quantities for age groups [0–14], [15–29], [30–39], and [40–49] for $ihr\_noise \sim N(1, 0.1); \psi_2 \sim N(0, 1)$. (Left) Estimated age-specific effective reproduction number, posterior mean estimate (dark blue line), and 95% credible intervals (light blue ribbon). (Middle) Observed daily age-specific SARS-CoV-2 hospitalization data (yellow barplot) versus posterior mean estimate (dark blue line) and 95% credible intervals (light blue ribbon). (Right) Observed daily age-specific SARS-CoV-2 case data (yellow barplot) versus posterior mean estimate (dark blue line) and 95% credible intervals (light blue ribbon).

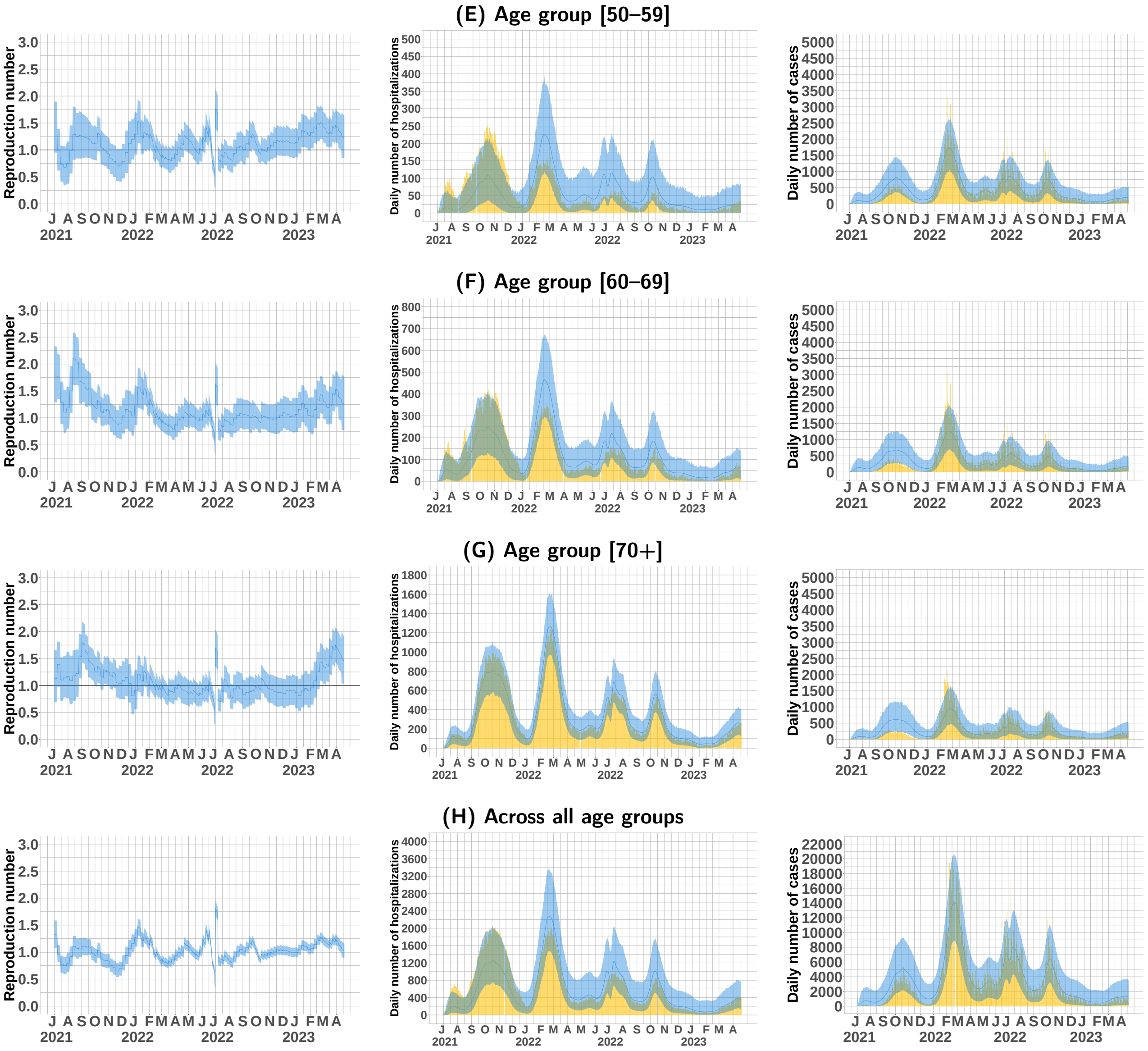


**Figure 2:** Model fits and key generated quantities for age groups [50–59], [60–69], [70+], and across all age groups for $ihr\_noise \sim N(1, 0.1); \psi_2 \sim N(0, 1)$. (Left) Estimated age-specific effective reproduction number, posterior mean estimate (dark blue line), and 95% credible intervals (light blue ribbon). (Middle) Observed daily age-specific SARS-CoV-2 hospitalization data (yellow barplot) versus posterior mean estimate (dark blue line) and 95% credible intervals (light blue ribbon). (Right) Observed daily age-specific SARS-CoV-2 case data (yellow barplot) versus posterior mean estimate (dark blue line) and 95% credible intervals (light blue ribbon).

#### 1.2 $ihr\_noise \sim N(1, 0.3); \psi_2 \sim N(0, 2)$

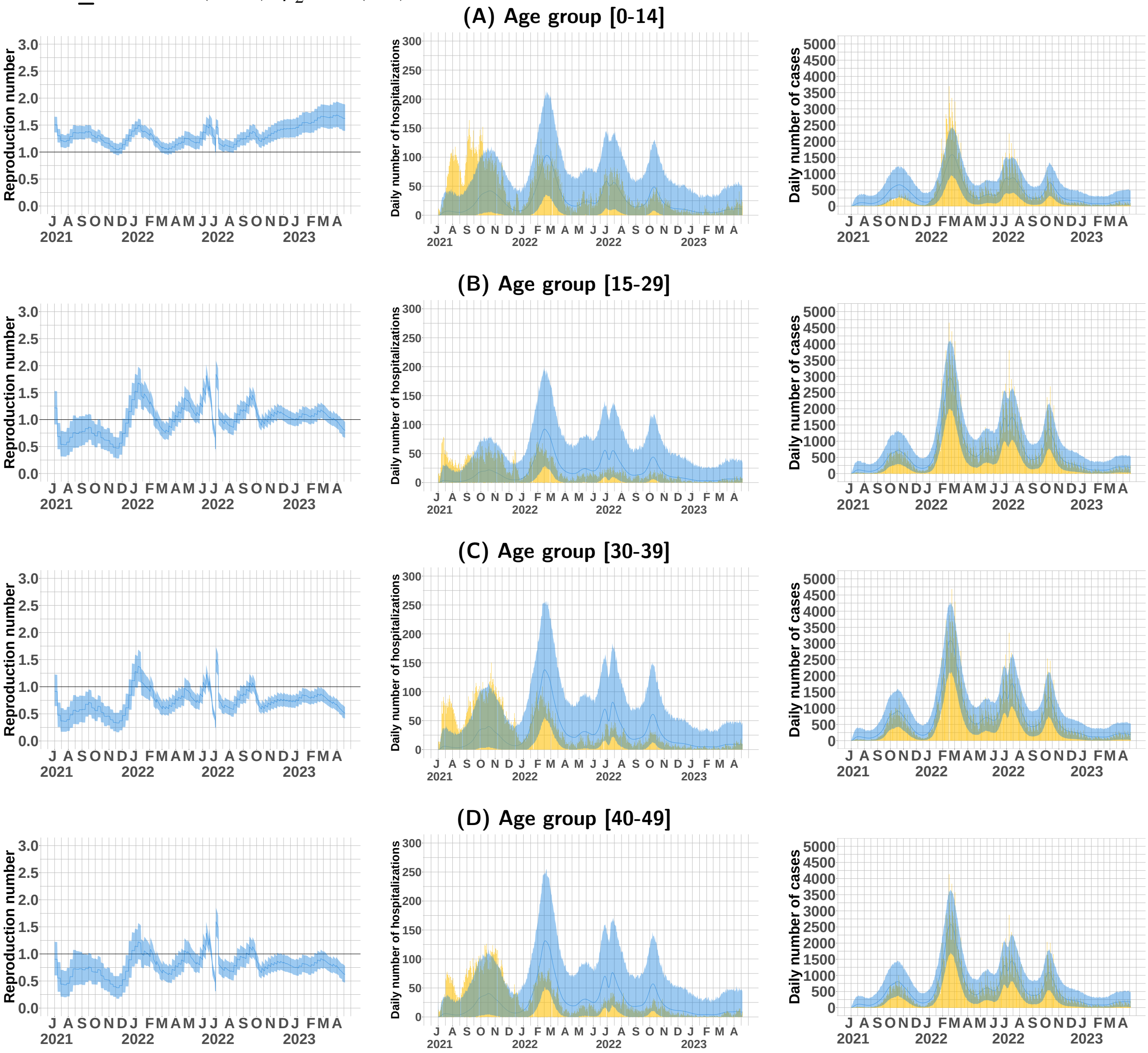


**Figure 3:** Model fits and key generated quantities for age groups [0–14], [15–29], [30–39], and [40–49] for $ihr\_noise \sim N(1, 0.3); \psi_2 \sim N(0, 2)$. (Left) Estimated age-specific effective reproduction number, posterior mean estimate (dark blue line), and 95% credible intervals (light blue ribbon). (Middle) Observed daily age-specific SARS-CoV-2 hospitalization data (yellow barplot) versus posterior mean estimate (dark blue line) and 95% credible intervals (light blue ribbon). (Right) Observed daily age-specific SARS-CoV-2 case data (yellow barplot) versus posterior mean estimate (dark blue line) and 95% credible intervals (light blue ribbon).

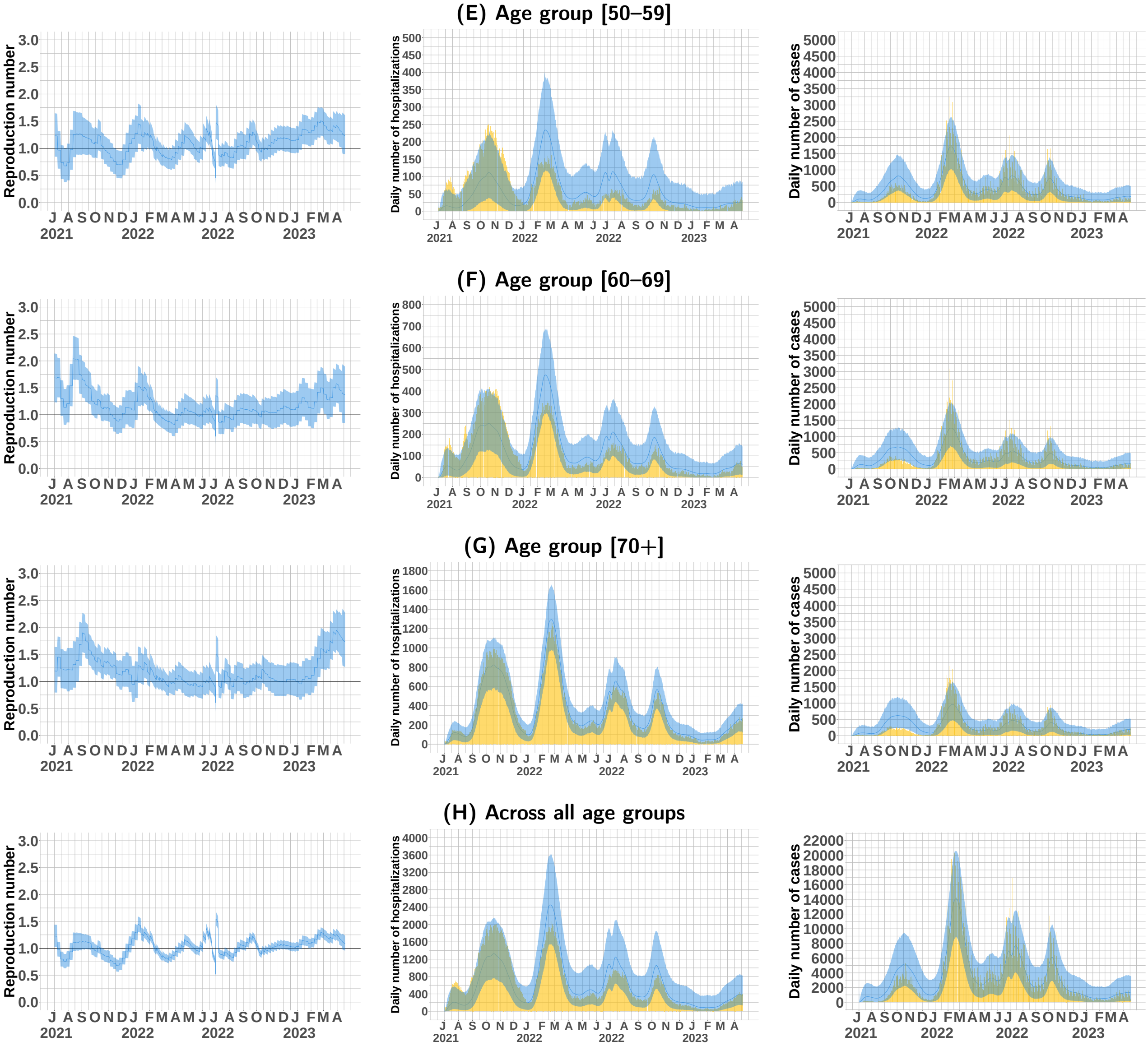


**Figure 4:** Model fits and key generated quantities for age groups [50–59], [60–69], [70+], and across all age groups for $ihr\_noise \sim N(1, 0.3); \psi_2 \sim N(0, 2)$. (Left) Estimated age-specific effective reproduction number, posterior mean estimate (dark blue line), and 95% credible intervals (light blue ribbon). (Middle) Observed daily age-specific SARS-CoV-2 hospitalization data (yellow barplot) versus posterior mean estimate (dark blue line) and 95% credible intervals (light blue ribbon). (Right) Observed daily age-specific SARS-CoV-2 case data (yellow barplot) versus posterior mean estimate (dark blue line) and 95% credible intervals (light blue ribbon).

##### 1.3 $ihr\_noise \sim N(1, 0.8); \psi_2 \sim N(0, 1)$

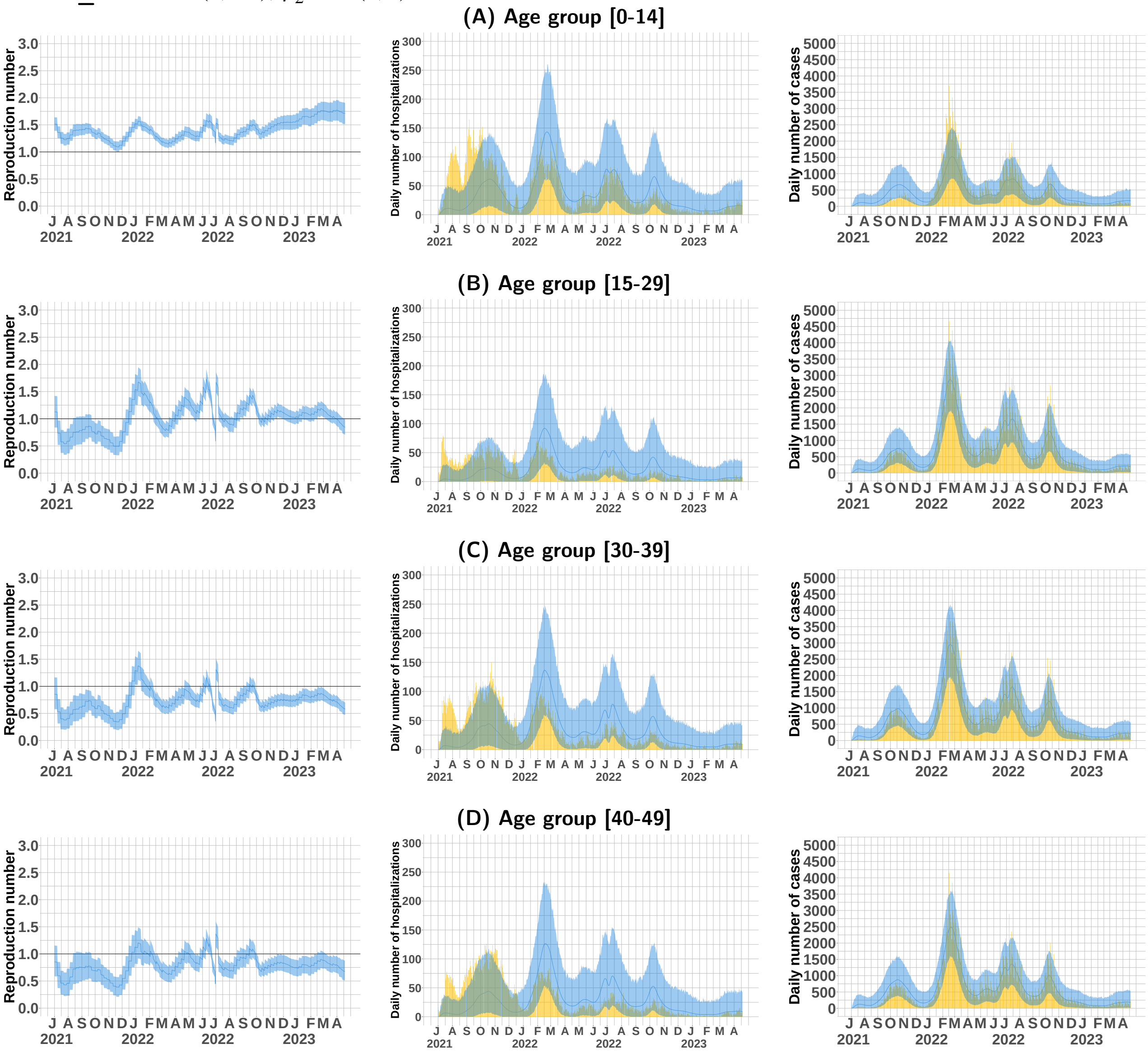


**Figure 5:** Model fits and key generated quantities for age groups [0–14], [15–29], [30–39], and [40–49] for $ihr\_noise \sim N(1, 0.8); \psi_2 \sim N(0, 1)$ . (Left) Estimated age-specific effective reproduction number, posterior mean estimate (dark blue line), and 95% credible intervals (light blue ribbon). (Middle) Observed daily age-specific SARS-CoV-2 hospitalization data (yellow barplot) versus posterior mean estimate (dark blue line) and 95% credible intervals (light blue ribbon). (Right) Observed daily age-specific SARS-CoV-2 case data (yellow barplot) versus posterior mean estimate (dark blue line) and 95% credible intervals (light blue ribbon).

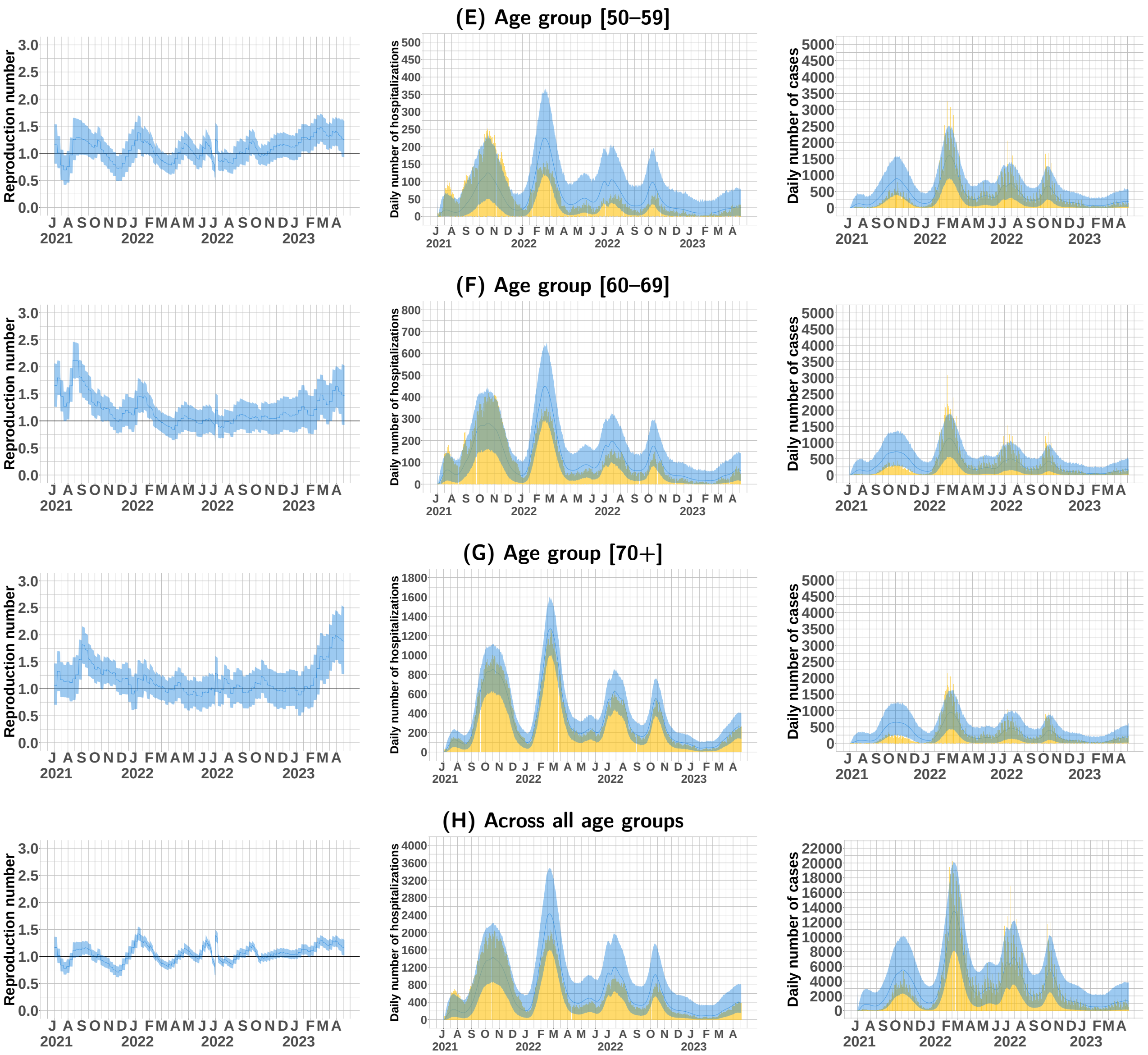


**Figure 6:** Model fits and key generated quantities for age groups [50–59], [60–69], [70+], and across all age groups for $ihr\_noise \sim N(1, 0.8); \psi_2 \sim N(0, 1)$. (Left) Estimated age-specific effective reproduction number, posterior mean estimate (dark blue line), and 95% credible intervals (light blue ribbon). (Middle) Observed daily age-specific SARS-CoV-2 hospitalization data (yellow barplot) versus posterior mean estimate (dark blue line) and 95% credible intervals (light blue ribbon). (Right) Observed daily age-specific SARS-CoV-2 case data (yellow barplot) versus posterior mean estimate (dark blue line) and 95% credible intervals (light blue ribbon).

##### 1.4 $ihr\_noise \sim N(1,1); \psi_2 \sim N(0,0.1)$

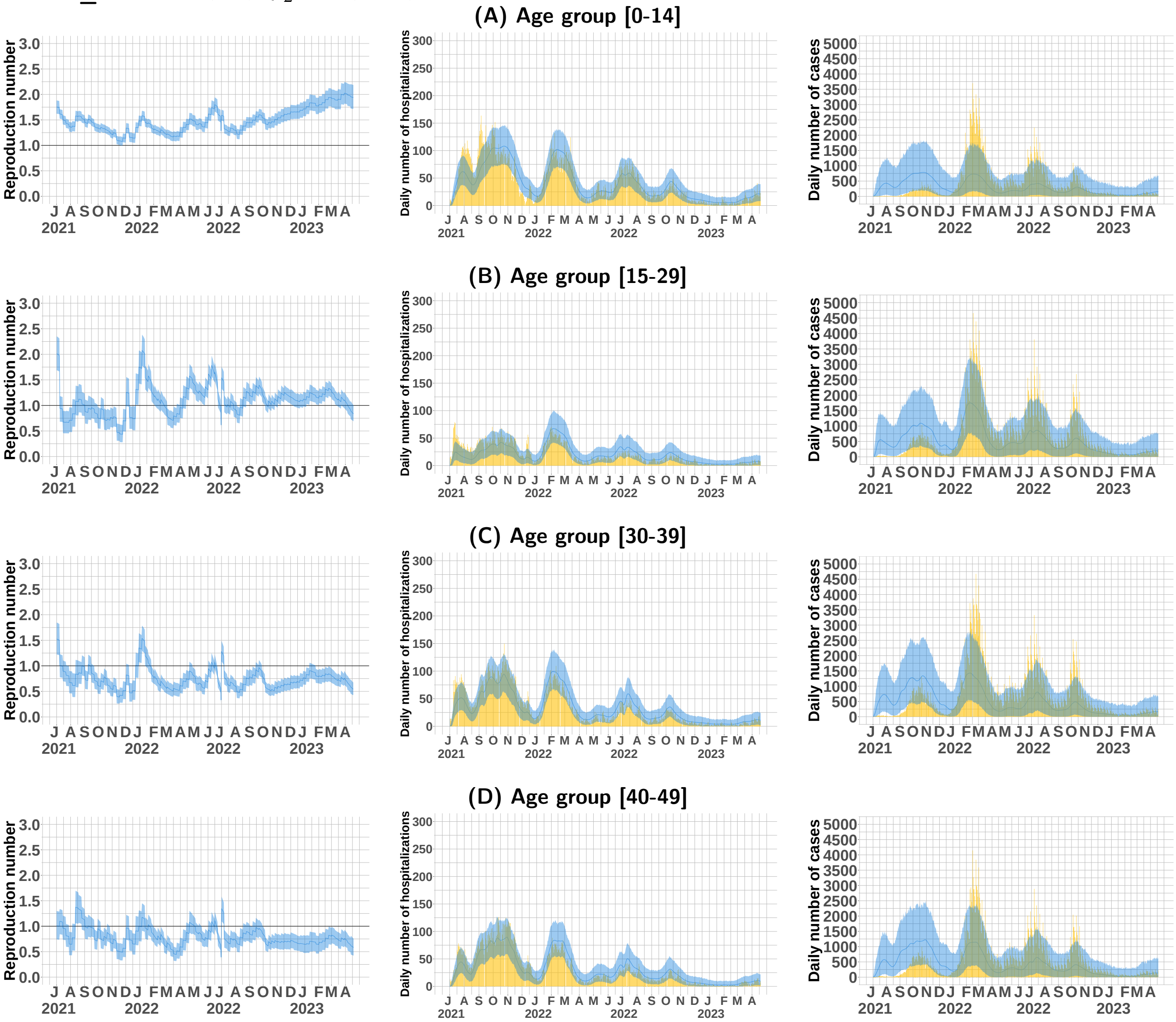


**Figure 7:** Model fits and key generated quantities for age groups [0–14], [15–29], [30–39], and [40–49] for $ihr\_noise \sim N(1,1); \psi_2 \sim N(0,0.1)$ . (Left) Estimated age-specific effective reproduction number, posterior mean estimate (dark blue line), and 95% credible intervals (light blue ribbon). (Middle) Observed daily age-specific SARS-CoV-2 hospitalization data (yellow barplot) versus posterior mean estimate (dark blue line) and 95% credible intervals (light blue ribbon). (Right) Observed daily age-specific SARS-CoV-2 case data (yellow barplot) versus posterior mean estimate (dark blue line) and 95% credible intervals (light blue ribbon).

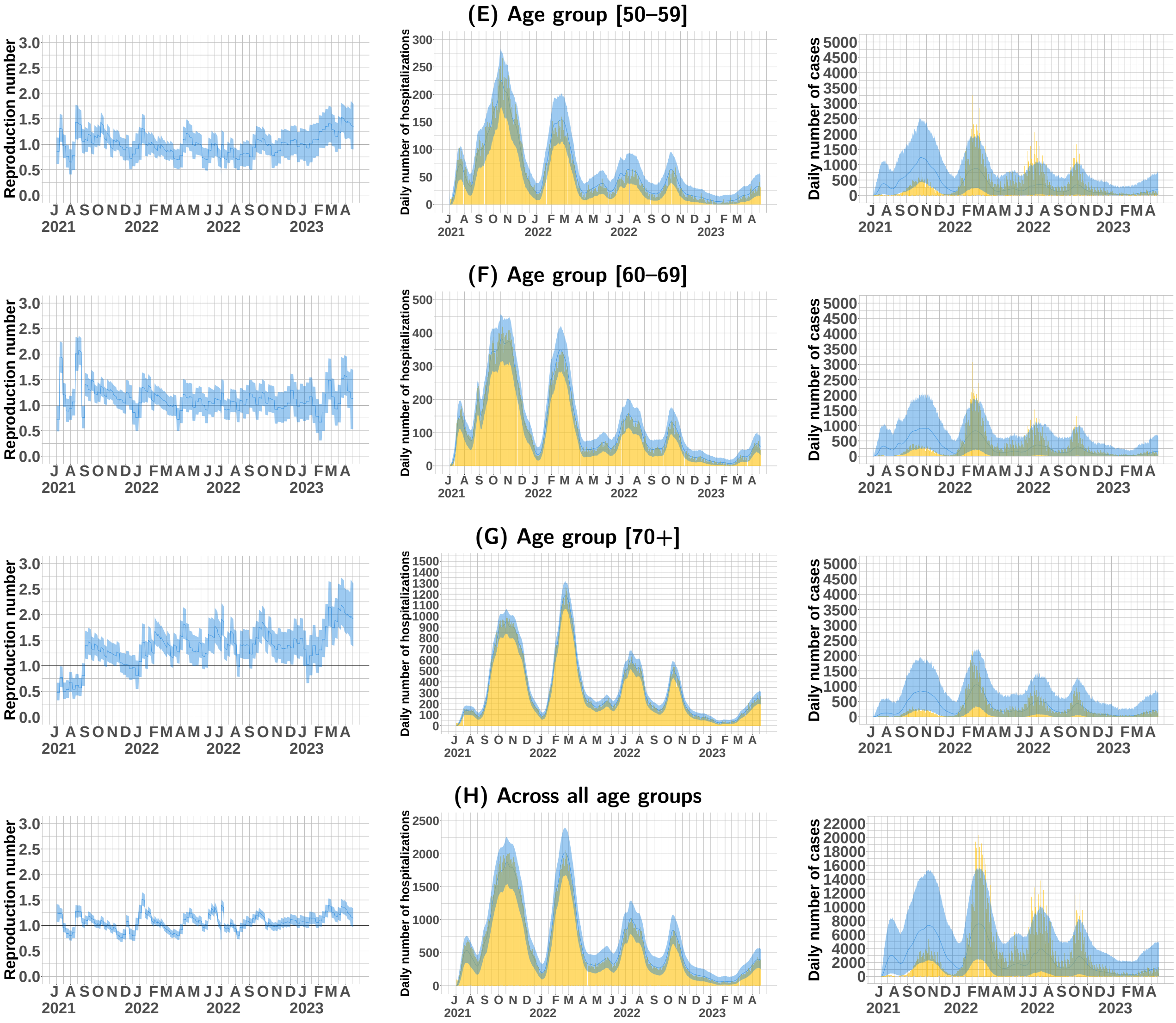


**Figure 8:** Model fits and key generated quantities for age groups [50–59], [60–69], [70+], and across all age groups for $ihr\_noise \sim N(1,1); \psi_2 \sim N(0,0.1)$. (Left) Estimated age-specific effective reproduction number, posterior mean estimate (dark blue line), and 95% credible intervals (light blue ribbon). (Middle) Observed daily age-specific SARS-CoV-2 hospitalization data (yellow barplot) versus posterior mean estimate (dark blue line) and 95% credible intervals (light blue ribbon). (Right) Observed daily age-specific SARS-CoV-2 case data (yellow barplot) versus posterior mean estimate (dark blue line) and 95% credible intervals (light blue ribbon).

#### 2. Elpd and ic for different priors of $ihr\_noise$ and $\psi_2$

| Settings (Standard deviation in $ihr\_noise$ and standard deviation in $\psi_2$) | elpd | ic |
|---|---|---|
| $ihr\_noise \sim N(1, 0.1); \psi_2 \sim N(0, 1)$ | -52050.5 | 104101.1 |
| $ihr\_noise \sim N(1, 0.3); \psi_2 \sim N(0, 2)$ | -50775.0 | 101550.0 |
| $ihr\_noise \sim N(1, 0.8); \psi_2 \sim N(0, 1)$ | -50359.8 | 100719.6 |
| $ihr\_noise \sim N(1, 1); \psi_2 \sim N(0, 0.1)$ | -51428.9 | 102857.8 |

**Table S2:** Elpd and ic for different priors of $ihr\_noise$ and $\psi_2$.

#### 3. Proportions of infections

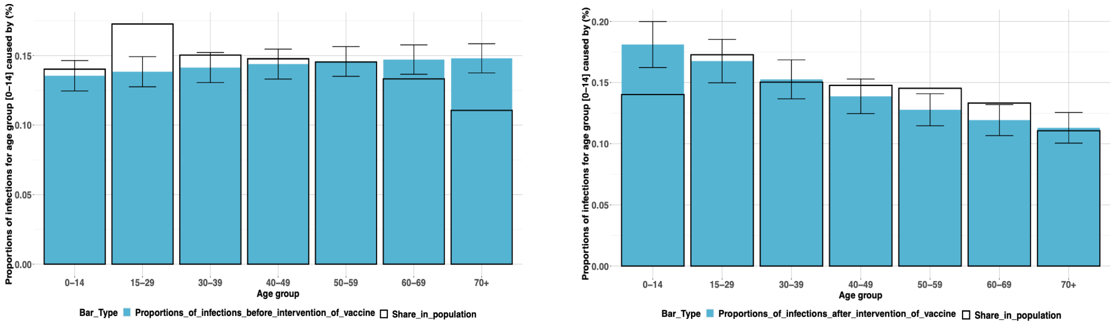


**Figure 9:** Comparison of SARS-CoV-2 proportions of infections for age group [0-14] before and after vaccine intervention in Singapore.

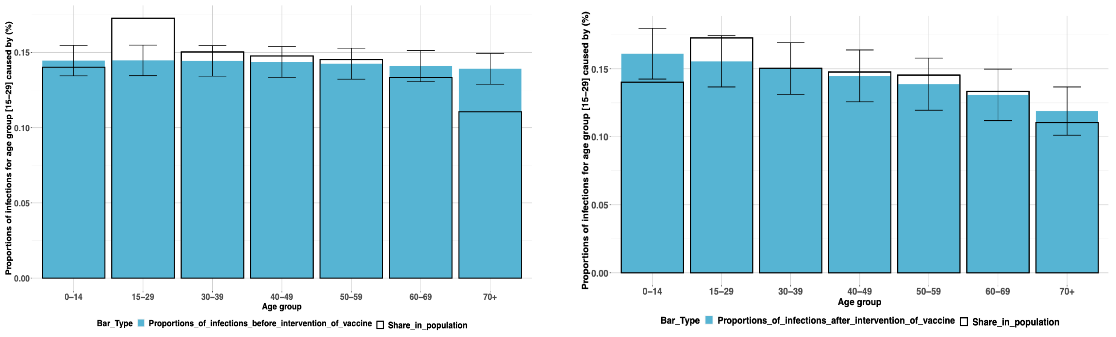


**Figure 10:** Comparison of SARS-CoV-2 proportions of infections for age group [15-29] before and after vaccine intervention in Singapore.

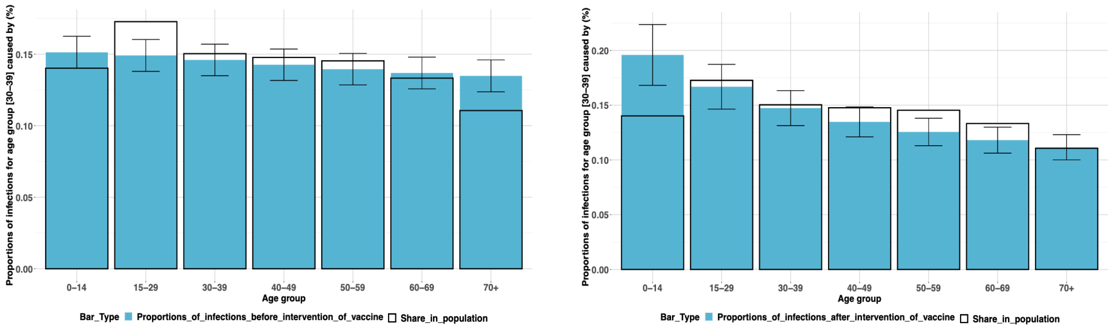


**Figure 11:** Comparison of SARS-CoV-2 proportions of infections for age group [30-39] before and after vaccine intervention in Singapore.

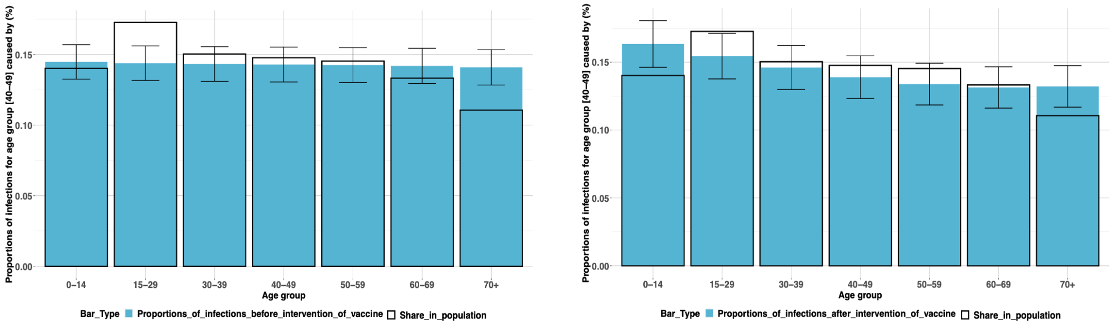


**Figure 12:** Comparison of SARS-CoV-2 proportions of infections for age group [40-49] before and after vaccine intervention in Singapore.

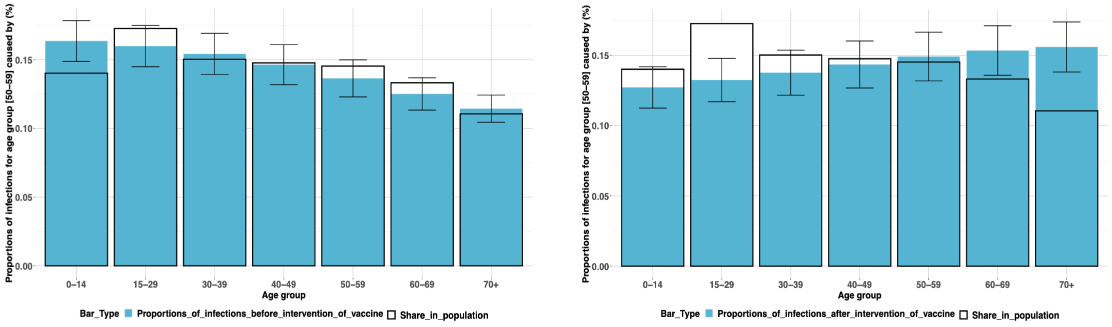


**Figure 13:** Comparison of SARS-CoV-2 proportions of infections for age group [50-59] before and after vaccine intervention in Singapore.

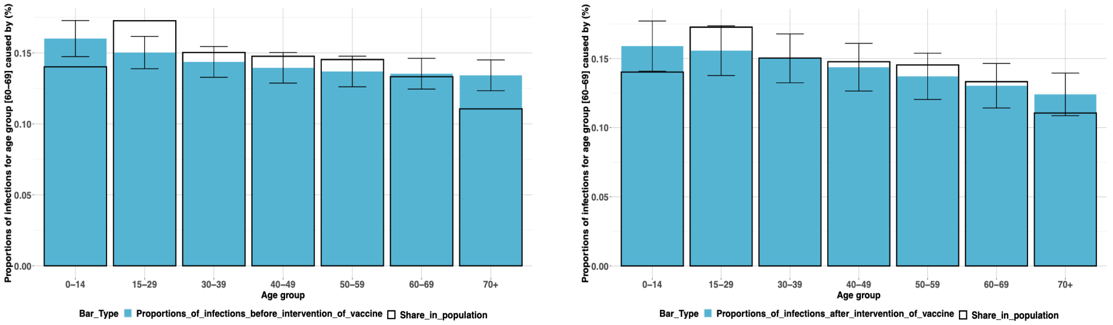


**Figure 14:** Comparison of SARS-CoV-2 proportions of infections for age group [60-69] before and after vaccine intervention in Singapore.

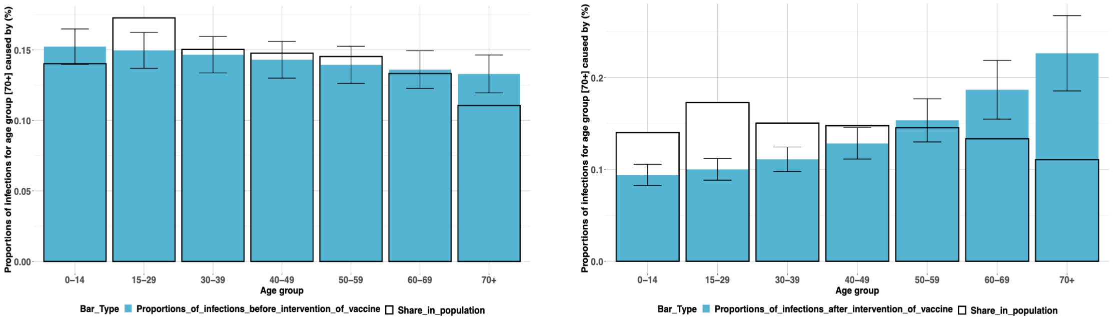


**Figure 15:** Comparison of SARS-CoV-2 proportions of infections for age group [70+] before and after vaccine intervention in Singapore.

### 4. Supplementary note of priors

From Monod et al. (2021), the transmission probability $\varrho_a$ is assumed to be

$$\varrho_a = \rho_0 \times \rho_a,$$

with $\rho_0 = R_0/AvgCntct$. We also assume $\log \rho_a$ for $a = 1, 2, \cdots, 7$, follows

$$\log \rho_1 = \log \rho_A,$$

$$\log \rho_{2:5} = rep(0, 4),$$

$$\log \rho_6 = \log \rho_B * 0.5,$$

$$\log \rho_7 = \log \rho_B,$$

with relative susceptibility parameters $\log \rho_A$ and $\log \rho_B$ following

$$\log \rho_A \sim N(-1.07, 0.22),$$

$$\log \rho_B \sim N(0.38, 0.16),$$

as specified in Monod et al. (2021).
